# Mortality among patients with severe traumatic brain injury: A systematic review and meta-analysis

**DOI:** 10.64898/2026.10.12.26365294

**Authors:** Frank Joshua Ruhazwe, Bienfait Mumbere, Mugenyi Michael, Theoneste Hakizimana, Owembabazi Elna

## Abstract

**Background:** Severe traumatic brain injury (TBI) is a leading cause of trauma death, yet reported mortality varies widely. We estimated the pooled mortality of severe TBI and explored sources of variation.

**Methods:** We searched PubMed/MEDLINE, Embase, Scopus, Web of Science, the Cochrane Library and Google Scholar from inception to January 2026 (PROSPERO CRD420261417287). Observational and interventional studies reporting the number of deaths among patients with severe TBI (Glasgow Coma Scale [GCS] ≤8) were eligible. Two reviewers independently selected studies, extracted data and assessed risk of bias with the Newcastle-Ottawa Scale. Logit-transformed proportions were pooled with a DerSimonian-Laird random-effects model; we report 95% confidence intervals (CI) and 95% prediction intervals (PI). Heterogeneity was explored by subgroup analysis, meta-regression and pre-specified sensitivity analyses, and certainty was rated with GRADE for prognosis.

**Results:** Twenty-four studies (11,544 patients; 5,234 deaths) from 15 countries, published 2001–2024, were included. Study-level mortality ranged from 18.2% to 71.1%. Pooled mortality was 40.9% (95% CI 35.7–46.3%; I² = 96.3%), with a 95% PI of 18.9–67.2%. Mortality differed by region (p = 0.002): 47.1% (37.6–56.8%) in Africa, 44.5% (40.2–48.8%) in Europe, 38.6% (25.4–53.7%) in Asia and 31.6% (26.5–37.2%) in the Americas. Mortality did not differ significantly by country income group, outcome time point, age group or publication year. Estimates were robust to alternative random-effects estimators, leave-one-out analysis (range 39.6–41.7%) and restriction to studies with independently verified counts (41.7%) or low risk of bias (38.4%). Certainty of evidence was low.

**Conclusions:** Approximately two in five patients with severe TBI die, but the wide prediction interval shows that the expected mortality in any single setting may lie anywhere between roughly one in five and two in three. Mortality was highest in African cohorts. Standardised definitions, outcome time points and reporting are needed to allow meaningful comparison across trauma systems.

## Introduction

Traumatic brain injury (TBI) is a major cause of death and disability worldwide, with an estimated 27 million new cases each year and a disproportionate burden in low- and middle-income countries (LMICs) [1–3]. Severe TBI, conventionally defined by a Glasgow Coma Scale (GCS) score of 3–8, accounts for a minority of cases but for most TBI-related deaths.

The management of severe TBI requires timely airway protection, neuroimaging, neurosurgical intervention and neurocritical care aimed at preventing secondary brain injury [4]. Access to each of these elements differs greatly between health systems.

Reported mortality after severe TBI varies several-fold between studies. Historical syntheses suggest that mortality fell substantially during the twentieth century before reaching a plateau, and that outcomes differ between geographical regions [5,6]. Large multinational datasets have shown worse outcomes in LMICs than in high-income countries (HICs) after adjustment for case mix, and prospective data on emergency neurosurgery show a similar gradient by level of human development [7–9].

These differences plausibly reflect prehospital care, time to definitive treatment, access to computed tomography, neurosurgery and intensive care, and treatment-limitation practices, as well as differences in case mix. Mortality estimates also depend on when death is measured. Early deaths within 24–48 hours largely reflect the primary injury, whereas deaths at 14 days, at hospital discharge, at 30 days or at 6 months also capture secondary brain injury, systemic complications and decisions about the continuation of care [10,11]. Established prognostic models therefore specify the outcome horizon explicitly [12,13].

An up-to-date synthesis that explicitly addresses between-study heterogeneity, rather than reporting a single pooled figure, would help clinicians, trauma-system planners and researchers to benchmark outcomes and design studies. We therefore conducted a systematic review and meta-analysis to estimate mortality among patients with severe TBI and to examine how it varies by geographical region, country income level, patient population and outcome time point.

## Materials and methods

### Protocol, registration and reporting

This review is reported in accordance with the Preferred Reporting Items for Systematic Reviews and Meta-Analyses (PRISMA) 2020 statement [14] (S1 Checklist) and follows methodological guidance for systematic reviews of observational studies reporting prevalence and cumulative incidence [15]. The protocol was registered prospectively in PROSPERO (CRD420261417287). Deviations from the protocol are listed at the end of this section.

### Information sources and search strategy

PubMed/MEDLINE, Embase, Scopus, Web of Science Core Collection and the Cochrane Library were searched from inception to January 2026. These searches were supplemented by Google Scholar (first 200 records per query) and by manual screening of the reference lists of included studies and relevant reviews.

Search strategies combined controlled vocabulary and free-text terms for traumatic brain injury and head injury, severity (severe, GCS, coma) and mortality or survival. The full strategy for each database is provided in S1 File.

### Eligibility criteria

#### Population

Patients of any age with severe TBI, defined as a GCS score ≤8 (equivalently, GCS <9 or GCS 3–8) at presentation or after resuscitation. Studies of mixed TBI severity were eligible only if the number of patients with severe TBI and the number of deaths among them could be extracted separately.

#### Outcome

All-cause mortality, defined as the number of deaths divided by the number of patients with severe TBI with known vital status. The outcome time point (≤48 hours, 7 days, 14 days, intensive care unit [ICU] or in-hospital, 30 days, or longer follow-up) was recorded as reported.

### Study designs

Prospective or retrospective cohort studies, registry or multicentre observational studies, and interventional studies reporting mortality for all enrolled patients with severe TBI.

### Exclusion criteria

Case reports, reviews, editorials, letters and conference abstracts without extractable data were excluded, as were studies of acquired brain injury that combined traumatic and non-traumatic aetiologies without separate TBI data. When several publications described the same or overlapping cohorts (same institution and overlapping recruitment period), the report with the largest sample and most complete outcome data was retained.

### Study selection

After removal of duplicates, two reviewers independently screened titles and abstracts and then assessed full texts against the eligibility criteria. Disagreements were resolved by discussion or by a third reviewer.

### Data extraction

Two reviewers independently extracted data using a piloted form: first author, publication year, country, geographical region (continent), World Bank income group, study design, setting and number of centres, recruitment period, population (age group and selection), definition of severe TBI, number of patients with severe TBI, number of deaths, mortality time point and duration of follow-up. Extracted numerators and denominators were then re-checked against the original publications.

Where a study reported mortality at more than one time point, the primary analysis used a single estimate per study, in the following order of preference: in-hospital (or ICU) mortality, 30-day mortality, 14-day mortality, then the longest reported follow-up. This ensured that no patient contributed more than once. Where outcome status was missing for some patients, the denominator was the number with known vital status.

### Risk-of-bias assessment

Two reviewers independently assessed risk of bias with the Newcastle-Ottawa Scale (NOS), adapted for single-group studies of prognosis [16]. Three domains were assessed:

- Selection: representativeness of the cohort (consecutive or population-based recruitment) and ascertainment of severe TBI.
- Comparability (adapted): whether the study described key prognostic case-mix variables, namely age, GCS, pupillary reactivity and computed tomography findings.
- Outcome: ascertainment of death, adequacy of follow-up and completeness of follow-up. Each domain and the overall judgement were rated as low, moderate or high risk of bias. Disagreements were resolved by consensus.

### Statistical analysis

#### Pooling of mortality

Study-level proportions with exact (Clopper-Pearson) 95% confidence intervals (CIs) were calculated. Proportions were logit-transformed and pooled using a DerSimonian-Laird random-effects model, and pooled estimates were back-transformed to the proportion scale [17,18]. The logit transformation was preferred over the Freeman-Tukey double-arcsine transformation because of known problems with back-transformation of the latter [19].

#### Heterogeneity

Heterogeneity was quantified with Cochran’s Q, I² and τ² [20]. A 95% prediction interval was calculated to describe the range within which the mortality of a new, comparable study population would be expected to lie [21].

#### Subgroup analyses and meta-regression

Pre-specified subgroup analyses were performed by mortality time point, geographical region (continent) and World Bank income group (HIC versus LMIC), with an additional post hoc subgroup by age group (paediatric, older adults, adult or all ages). Differences between subgroups were tested with a Q test for between-group heterogeneity based on separate within-subgroup τ² [22]. Random-effects meta-regression (method-of-moments τ²) was used to examine publication year and log sample size as continuous moderators.

#### Sensitivity analyses

The robustness of the pooled estimate was assessed using alternative statistical methods and restricted study sets:

- Alternative methods: restricted maximum likelihood (REML) estimation of τ²; the Hartung-Knapp-Sidik-Jonkman (HKSJ) confidence interval [23]; the Freeman-Tukey transformation; and a fixed-effect model.
- Influence analyses: leave-one-out analysis and screening for outliers using standardised deleted residuals [24].
- Restricted analyses: studies with low overall risk of bias; studies whose counts were confirmed against the full publication; studies reporting in-hospital or ICU mortality; unselected populations with confirmed severe TBI; studies with ≥50 patients; and adult or all-age cohorts.
- analysis of unselected populations excluded studies restricted to survivors beyond day 5, to operated older adults or to prehospital-intubation cohorts, and the study that included trauma patients with GCS ≤8 without confirmed TBI.

#### Small-study effects

Small-study effects were explored with a funnel plot and Egger’s regression test [25]. Results were interpreted cautiously, because funnel-plot methods perform poorly for meta-analyses of proportions with high heterogeneity [26].

#### Software

Two-sided p < 0.05 was considered statistically significant. Analyses were performed in Python (version 3; NumPy and SciPy). The analysis code and dataset are provided as Supporting Information (S1 Dataset, S1 Code).

#### Certainty of evidence

Certainty in the pooled mortality estimate was rated with the GRADE approach for prognostic evidence about event rates [27]. Because the evidence came from cohort studies of representative patients, the rating started at high and was downgraded for risk of bias, inconsistency, indirectness, imprecision and publication bias as appropriate.

#### Deviations from the protocol

The following were not specified in the registered protocol: the hierarchy for selecting one mortality estimate per study; the prediction interval; the HKSJ and REML sensitivity analyses; the subgroups by income group and age group; the meta-regression; and the sensitivity analyses restricted to studies with confirmed counts or unselected populations. These were added to address heterogeneity and data quality and did not alter the review question.

## Results

### Study selection

The searches identified 8,964 records. After removal of 2,781 duplicates, 6,183 titles and abstracts were screened and 191 full-text reports were assessed for eligibility. Of these, 167 were excluded, including one study of severe acquired brain injury of mixed traumatic and non-traumatic aetiology in a rehabilitation setting [28]. Twenty-four studies were included in the review and meta-analysis [29–52] (Fig 1).

**Fig 1.**
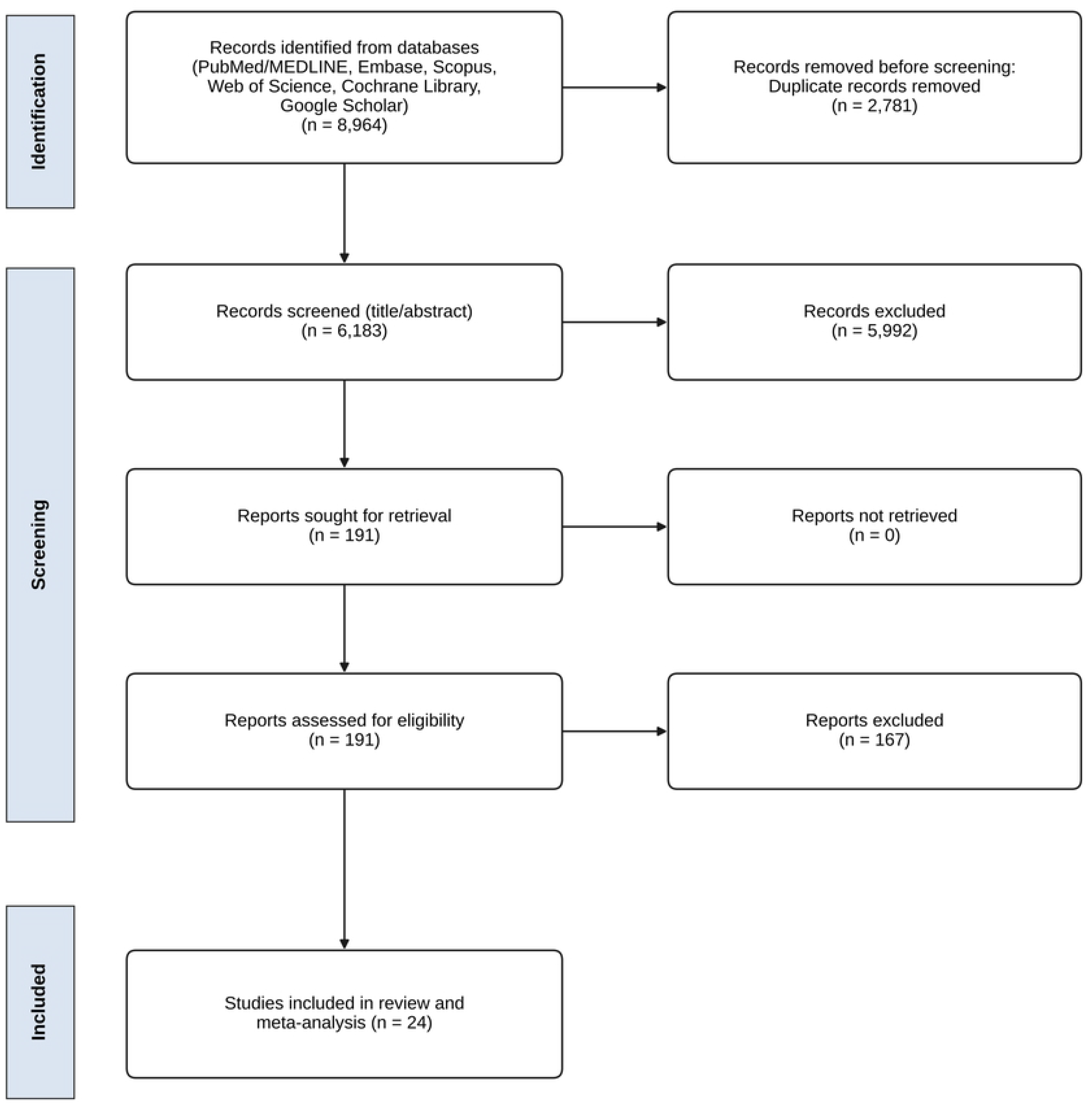
PRISMA 2020 flow diagram of study identification, screening and inclusion.

### Characteristics of included studies

The 24 studies were published between 2001 and 2024 and included 11,544 patients with severe TBI, of whom 5,234 died (Table 1). Sample sizes ranged from 11 to 3,476 patients.

**Table 1.** Characteristics of the 24 included studies.

| **Study** | **Country** | **Design** | **Population** | **sTBI definition** | **Mortality time point** | **Deaths/N** | **Mortality, % (95% CI)** |
| --- | --- | --- | --- | --- | --- | --- | --- |
| Mwang'ombe & Kiboi 2001 [29] | Kenya | Retrospective cohort | sTBI, all ages | GCS $\leq 8$ | In-hospital | 381/677 | 56.3 (52.4–60.1) |
| Jiang et al. 2002 [30] | China | Cohort | sTBI, all ages | GCS $\leq 8$ | 1 year | 249/846 | 29.4 (26.4–32.6) |
| Davis et al. 2003 [31] | USA | Prospective trial cohort with matched historical controls | sTBI, prehospital RSI cohort + matched controls | GCS 3–8 | In-hospital | 221/836 | 26.4 (23.5–29.6) |
| Jeremitsky et al. 2005 [32] | USA | Retrospective cohort | sTBI surviving >5 days | GCS $\leq 8$ | In-hospital | 24/77 | 31.2 (21.1–42.7) |
| Boto et al. 2006 [33] | Spain | Prospective database cohort | sTBI, age $\geq 15$ y | GCS $\leq 8$ after resuscitation | End of follow-up | 419/895 | 46.8 (43.5–50.1) |
| Opondo & Mwang'ombe 2007 [34] | Kenya | Prospective cohort | Adult sTBI admitted to ICU | GCS 3–8 | ICU/in-hospital | 46/87 | 52.9 (41.9–63.7) |
| Seyed Saadat et al. 2012 [35] | Iran | Cohort | Paediatric sTBI (PICU) | GCS $\leq 8$ | In-hospital | 49/122 | 40.2 (31.4–49.4) |
| Rahimi et al. 2014 [36] | Iran | Cohort | Paediatric sTBI | GCS $\leq 8$ | In-hospital | 34/108 | 31.5 (22.9–41.1) |
| Dawes et al. 2015 [37] | USA | Retrospective multicentre cohort | sTBI, adults | GCS $\leq 8$ | In-hospital | 319/822 | 38.8 (35.5–42.2) |
| Tran et al. 2015 [38] | Uganda | Retrospective cohort | sTBI, all ages | GCS $\leq 8$ | In-hospital | 31/120 | 25.8 (18.3–34.6) |

| Study | Country | Design | Population | sTBI definition | Mortality time point | Deaths/N | Mortality, % (95% CI) |
| --- | --- | --- | --- | --- | --- | --- | --- |
| Jerome et al. 2017 [39] | South Africa | Retrospective registry audit | sTBI, all ages | GCS <8 | In-hospital | 110/329 | 33.4 (28.4–38.8) |
| Krebs et al. 2017 [40] | Rwanda | Prospective cohort | sTBI subgroup of TBI registry | GCS 3–8 | In-hospital | 37/66 | 56.1 (43.3–68.3) |
| Smart et al. 2017 [41] | Tanzania | Prospective cohort | sTBI, all ages | GCS ≤8 | 2 weeks | 40/115 | 34.8 (26.1–44.2) |
| Areas et al. 2019 [42] | Brazil | Prospective multicentre cohort | sTBI, adults | GCS ≤8 after resuscitation | In-hospital | 83/266 | 31.2 (25.7–37.1) |
| Rodríguez-Triviño et al. 2019 [43] | Colombia | Cohort | sTBI, adults | GCS ≤8 | In-hospital | 26/83 | 31.3 (21.6–42.4) |
| Kumar et al. 2020 [44] | USA | Prospective cohort | sTBI, adults | GCS ≤8 | In-hospital | 48/157 | 30.6 (23.5–38.4) |
| Basha et al. 2021 [45] | Egypt | Retrospective cohort | sTBI, single centre | GCS ≤8 | Not stated† | 8/20 | 40.0 (19.1–63.9) |
| Bossers et al. 2021 [46] | Netherlands | Prospective multicentre observational | Prehospital GCS ≤8, confirmed TBI (head AIS ≥3) | Prehospital GCS ≤8 | 30 days | 656/1,547 | 42.4 (39.9–44.9) |
| Mangat et al. 2021 [47] | Tanzania | Prospective cohort | sTBI, adults | GCS ≤8 within 24 h | 2 weeks | 290/433 | 67.0 (62.3–71.4) |
| Matovu et al. 2021 [48] | Uganda | Prospective cohort | sTBI, all ages | GCS ≤8 | 30 days | 47/99 | 47.5 (37.3–57.8) |
| Demlie et al. 2023 [49] | Ethiopia | Retrospective multicentre cohort | sTBI subgroup of TBI cohort | GCS ≤8 | In-hospital | 59/83 | 71.1 (60.1–80.5) |
| Shen et al. 2023 [50] | China | Retrospective cohort | Older adults with sTBI undergoing surgery | GCS ≤8 | In-hospital | 98/269 | 36.4 (30.7–42.5) |
| Dibera et al. 2024 [51] | Ethiopia | Prospective cohort | sTBI subgroup admitted to surgical ward | GCS $\leq 8$ | In-hospital | 2/11 | 18.2 (2.3–51.8) |
| Soni et al. 2024 [52] | India | Prospective multicentre registry cohort | Trauma patients with GCS $\leq 8$ (TBI not confirmed in all) | GCS $\leq 8$ | In-hospital (30-day) | 1,957/3,476 | 56.3 (54.6–58.0) |
| **Total (24 studies)** |  |  |  |  |  | **5,234/11,544** |  |

AIS, Abbreviated Injury Scale; CI, exact (Clopper-Pearson) 95% confidence interval; GCS, Glasgow Coma Scale; ICU, intensive care unit; RSI, rapid sequence intubation; sTBI, severe traumatic brain injury. Bossers et al. = BRAIN-PROTECT study; denominator restricted to patients with confirmed TBI (head AIS $\geq 3$ ) and known 30-day status. Mangat et al.: denominator excludes 29 patients with unknown outcome. Soni et al. enrolled trauma patients with GCS $\leq 8$ without confirmation of TBI in all patients. †Mortality window not explicitly stated; grouped with in-hospital/ICU studies in subgroup analyses.

### Geographical distribution

Eleven studies were conducted in Africa (Kenya, Uganda, Tanzania, Rwanda, Ethiopia, Egypt and South Africa), five in Asia (China, Iran and India), two in Europe (Spain and the Netherlands) and six in the Americas (United States, Colombia and Brazil). Eighteen studies were from LMICs and six from HICs.

Cohorts from the same institutions (Kenyatta National Hospital, Kenya [29,34]; Mulago National Referral Hospital, Uganda [38,48]) had non-overlapping recruitment periods, and the two Tanzanian cohorts were from different hospitals [41,47].

### Populations and outcome definitions

Two studies were restricted to children and one to older adults undergoing surgery. One study enrolled all trauma patients with GCS ≤8, not all of whom had confirmed TBI [52], and one was restricted to patients surviving more than five days [32].

Eighteen studies reported in-hospital or ICU mortality (including one in which the time window was not explicitly stated [45]), two reported 14-day mortality, two 30-day mortality and two mortality at longer follow-up.

### Risk of bias in included studies

Overall risk of bias was low in 8 studies (33%) and moderate in 16 (67%); no study was rated at high risk (Fig 2; Table 2). Selection was at low risk in 16 studies (67%) and outcome assessment in 23 (96%).

**Fig 2.**
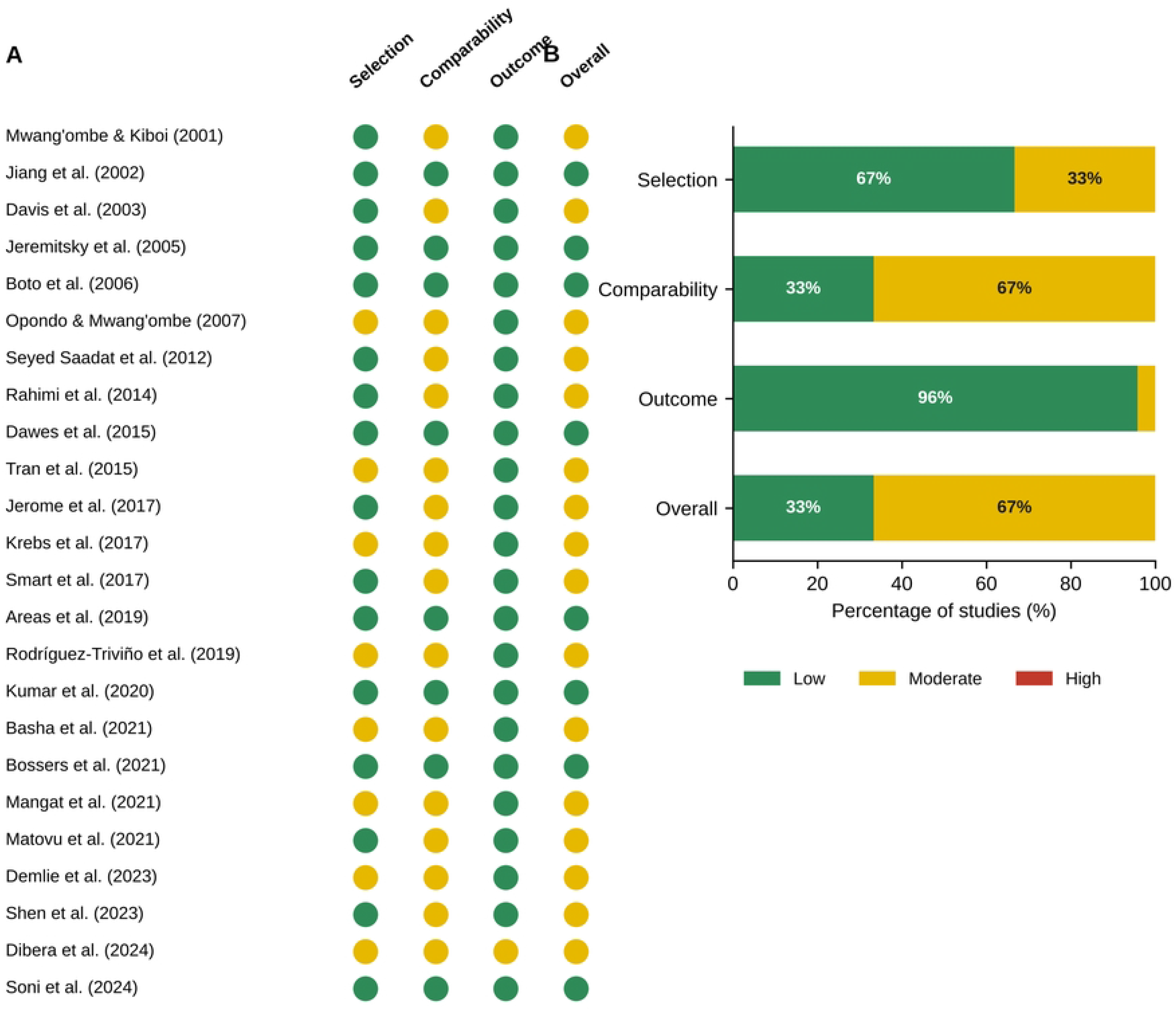
Risk-of-bias assessment of included studies (adapted Newcastle-Ottawa Scale). (A) Judgements by study and domain. (B) Proportion of studies at each level of risk by domain.

**Table 2.** Risk-of-bias assessment of included studies (adapted Newcastle-Ottawa Scale).

| Study | Selection | Comparability | Outcome | Overall |
| --- | --- | --- | --- | --- |
| Mwang'ombe & Kiboi 2001 | Low | Moderate | Low | Moderate |
| Jiang et al. 2002 | Low | Low | Low | Low |
| Davis et al. 2003 | Low | Moderate | Low | Moderate |
| Jeremitsky et al. 2005 | Low | Low | Low | Low |
| Boto et al. 2006 | Low | Low | Low | Low |
| Opondo & Mwang'ombe 2007 | Moderate | Moderate | Low | Moderate |
| Seyed Saadat et al. 2012 | Low | Moderate | Low | Moderate |
| Rahimi et al. 2014 | Low | Moderate | Low | Moderate |
| Dawes et al. 2015 | Low | Low | Low | Low |
| Tran et al. 2015 | Moderate | Moderate | Low | Moderate |
| Jerome et al. 2017 | Low | Moderate | Low | Moderate |
| Krebs et al. 2017 | Moderate | Moderate | Low | Moderate |
| Smart et al. 2017 | Low | Moderate | Low | Moderate |
| Areas et al. 2019 | Low | Low | Low | Low |
| Rodríguez-Triviño et al. 2019 | Moderate | Moderate | Low | Moderate |
| Kumar et al. 2020 | Low | Low | Low | Low |
| Basha et al. 2021 | Moderate | Moderate | Low | Moderate |
| Bossers et al. 2021 | Low | Low | Low | Low |
| Mangat et al. 2021 | Moderate | Moderate | Low | Moderate |
| Matovu et al. 2021 | Low | Moderate | Low | Moderate |
| Demlie et al. 2023 | Moderate | Moderate | Low | Moderate |
| Shen et al. 2023 | Low | Moderate | Low | Moderate |
| Dibera et al. 2024 | Moderate | Moderate | Moderate | Moderate |
| Soni et al. 2024 | Low | Low | Low | Low |

Overall risk was low in 8/24 (33%) and moderate in 16/24 (67%) studies.

The main limitation was comparability: only 8 studies (33%) adequately described the key prognostic case-mix variables, and several single-centre studies used convenience samples or retrospective record review.

### Overall pooled mortality

Mortality ranged from 18.2% (2/11) [51] to 71.1% (59/83) [49]. The pooled mortality was 40.9% (95% CI 35.7–46.3%), with considerable heterogeneity (I² = 96.3%; τ² = 0.261; Q = 624.8, df = 23, p < 0.001). The 95% prediction interval was 18.9–67.2% (Fig 3).

**Fig 3.**
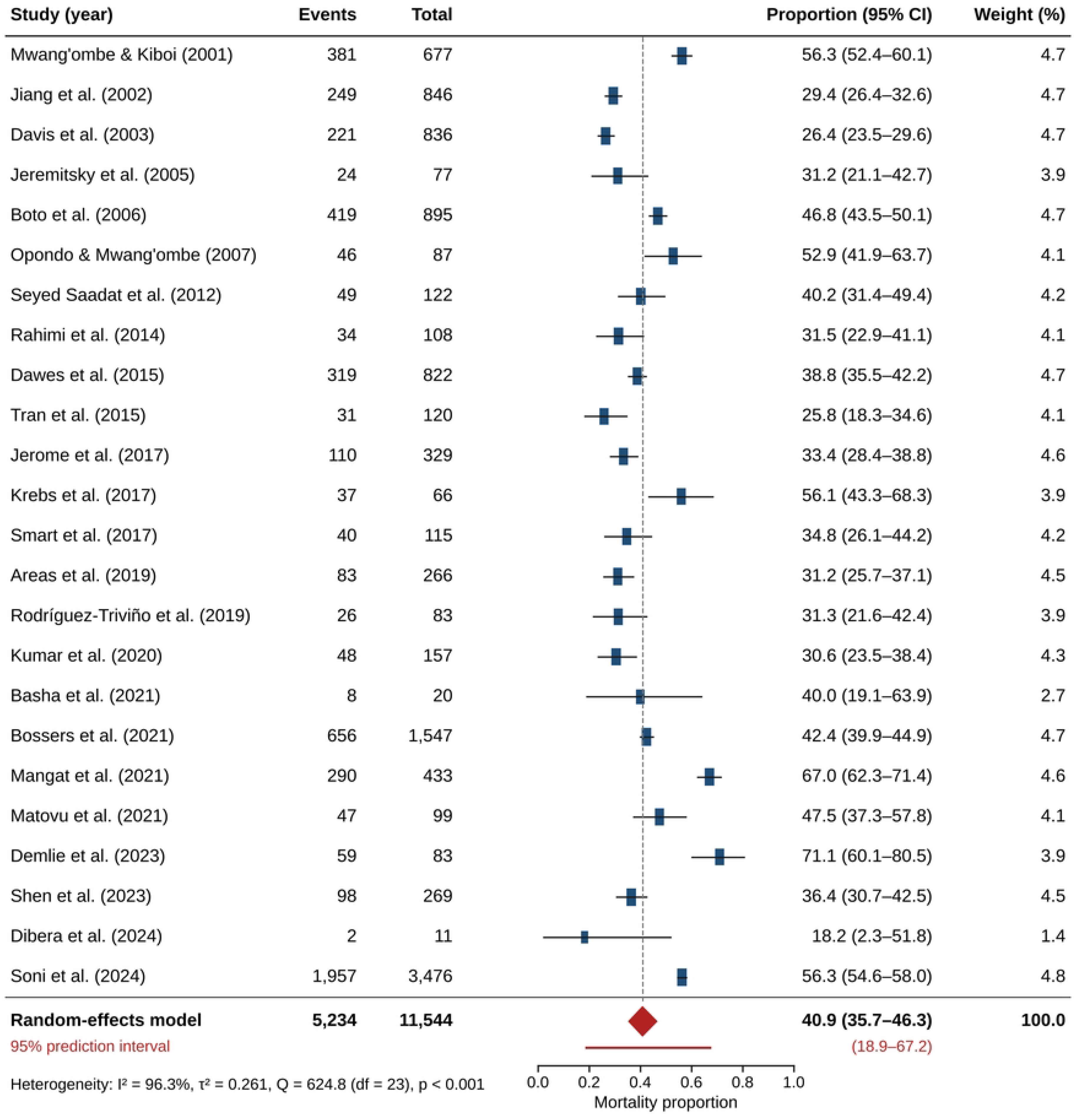
Forest plot of mortality among patients with severe traumatic brain injury. Squares show study estimates (size proportional to random-effects weight) with exact 95% confidence intervals; the diamond shows the pooled random-effects estimate and the red line the 95% prediction interval.

### Subgroup analyses and meta-regression

Results of all subgroup analyses are summarised in Table 3.

**Table 3.** Subgroup analyses of mortality among patients with severe traumatic brain injury.

| Subgroup | Studies | Patients | Deaths | Pooled mortality, % (95% CI) | I<sup>2</sup> (%) | p for difference |
| --- | --- | --- | --- | --- | --- | --- |
| **Mortality time point** |  |  |  |  |  | **0.728** |
| In-hospital or ICU | 18 | 7,609 | 3,533 | 39.5 (32.9–46.4) | 96.2 |  |
| 14 days | 2 | 548 | 330 | 51.2 (22.1–79.6) | 97.3 |  |
| 30 days | 2 | 1,646 | 703 | 42.7 (40.3–45.1) | 0.0 |  |
| >30 days / end of follow-up | 2 | 1,741 | 668 | 37.7 (22.6–55.8) | 98.2 |  |
| **Geographical region** |  |  |  |  |  | **0.002** |
| Africa | 11 | 2,040 | 1,051 | 47.1 (37.6–56.8) | 93.2 |  |
| Asia | 5 | 4,821 | 2,387 | 38.6 (25.4–53.7) | 98.2 |  |
| Europe | 2 | 2,442 | 1,075 | 44.5 (40.2–48.8) | 77.6 |  |
| Americas | 6 | 2,241 | 721 | 31.6 (26.5–37.2) | 82.9 |  |

| Subgroup | Studies | Patients | Deaths | Pooled mortality,<br>% (95% CI) | I<sup>2</sup><br>(%) | p for<br>difference |
| --- | --- | --- | --- | --- | --- | --- |
| **Country income group** |  |  |  |  |  | **0.192** |
| Low- and middle-income | 18 | 7,210 | 3,547 | 42.7 (35.8–49.8) | 96.0 |  |
| High-income | 6 | 4,334 | 1,687 | 36.2 (29.9–43.1) | 94.5 |  |
| **Age group** |  |  |  |  |  | **0.378** |
| Adults or all ages | 21 | 11,045 | 5,053 | 41.6 (36.0–47.5) | 96.7 |  |
| Paediatric | 2 | 230 | 83 | 36.0 (27.9–44.9) | 46.4 |  |
| Older adults<br>(operated) | 1 | 269 | 98 | 36.4 (30.9–42.4) | – |  |
| **Overall** | **24** | **11,544** | **5,234** | **40.9 (35.7–46.3)** | **96.3** |  |

Random-effects (DerSimonian-Laird) estimates on the logit scale. p values from the Q test for between-subgroup heterogeneity. ICU, intensive care unit.

### Geographical region

Pooled mortality differed by region (test for subgroup differences p = 0.002). It was highest in Africa (47.1%, 95% CI 37.6–56.8%; 11 studies) and lowest in the Americas (31.6%, 26.5– 37.2%; 6 studies), with intermediate estimates for Europe (44.5%, 40.2–48.8%; 2 studies) and Asia (38.6%, 25.4–53.7%; 5 studies) (Fig 4).

**Fig 4.**
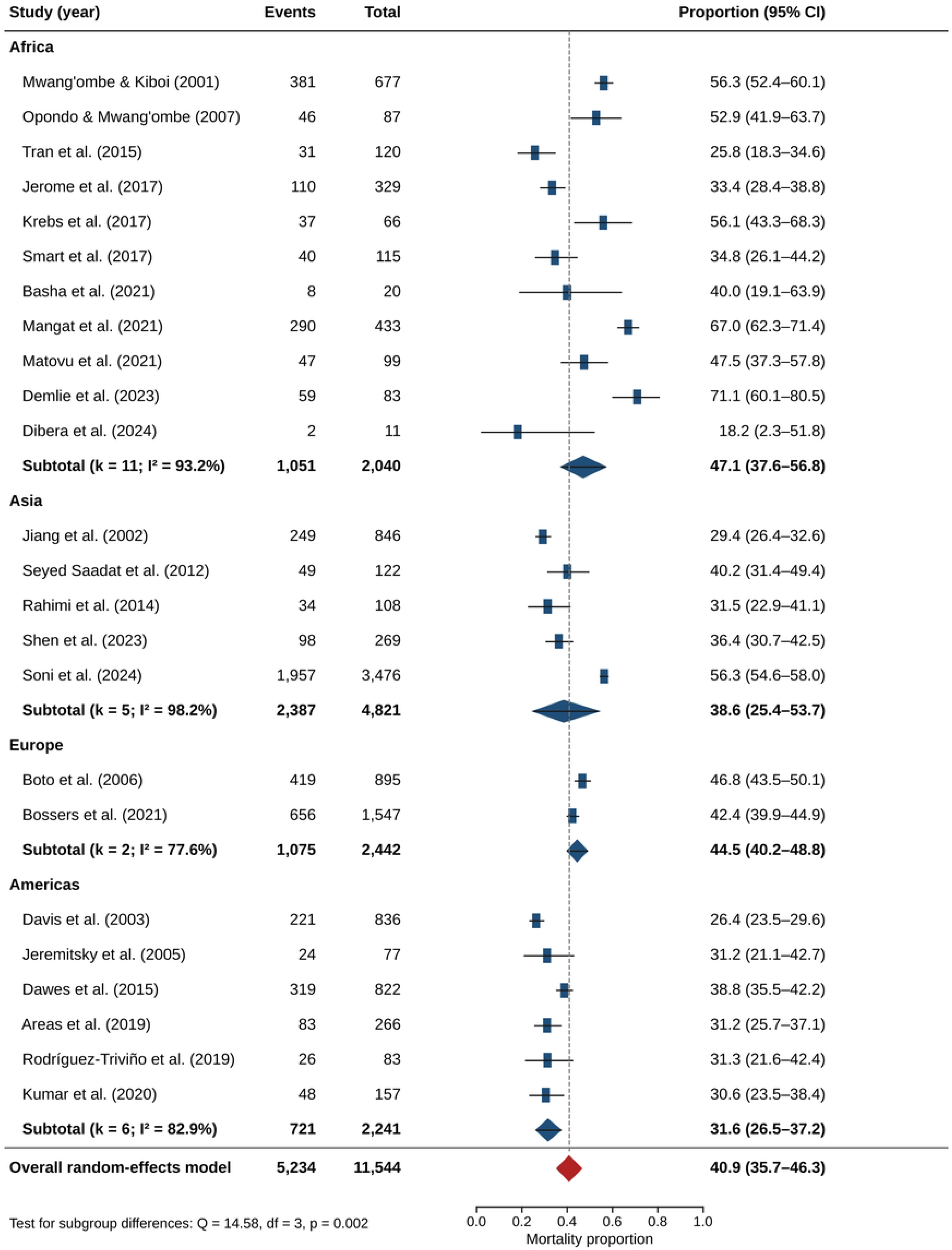
Forest plot of mortality among patients with severe traumatic brain injury by geographical region.

### Country income group

Mortality was numerically higher in LMICs (42.7%, 35.8–49.8%) than in HICs (36.2%, 29.9– 43.1%), but the difference was not statistically significant (p = 0.19).

### Mortality time point

Pooled in-hospital or ICU mortality was 39.5% (32.9–46.4%; 18 studies). Pooled 14-day mortality was 51.2% (22.1–79.6%), 30-day mortality 42.7% (40.3–45.1%) and mortality at longer follow-up 37.7% (22.6–55.8%), each from only two studies (Fig 5). There was no statistically significant difference between time points (p = 0.73), and the small number of studies outside the in-hospital category precluded a reliable assessment of a time gradient.

**Fig 5.**
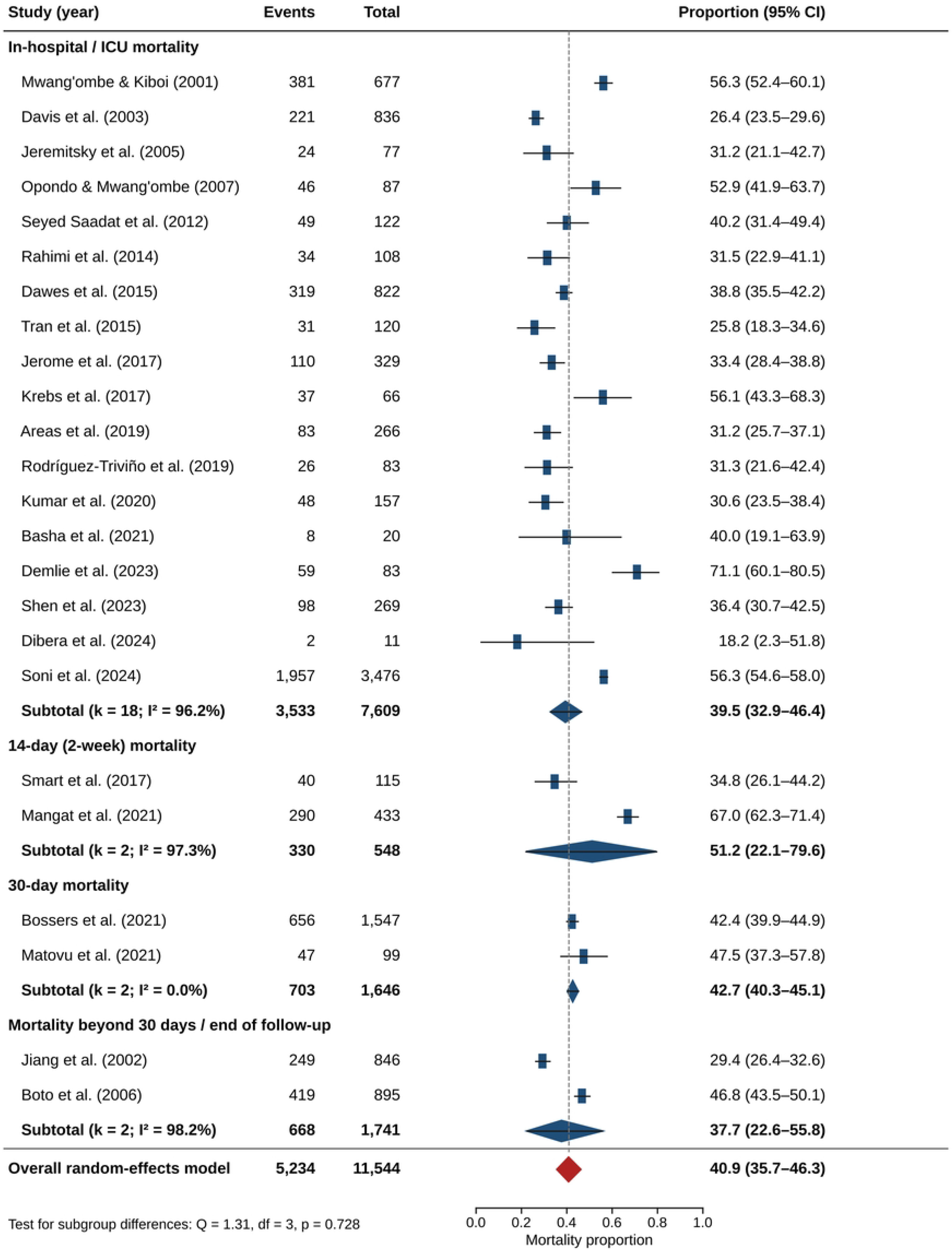
Forest plot of mortality among patients with severe traumatic brain injury by mortality time point.

### Age group and meta-regression

Mortality did not differ by age group (p = 0.38). In meta-regression, neither publication year (change in logit mortality per decade 0.15, standard error 0.15; p = 0.32) nor log sample size (p = 0.44) was associated with mortality.

### Sensitivity analyses

#### Alternative statistical methods

The pooled estimate was unchanged with REML (40.9%, 35.6–46.3%), with the HKSJ interval (40.9%, 35.3–46.7%) and with the Freeman-Tukey transformation (40.9%, 35.6–46.4%). The fixed-effect estimate was higher (45.7%, 44.8–46.6%) because of the weight given to the largest study [52] (Table 4).

**Table 4.**
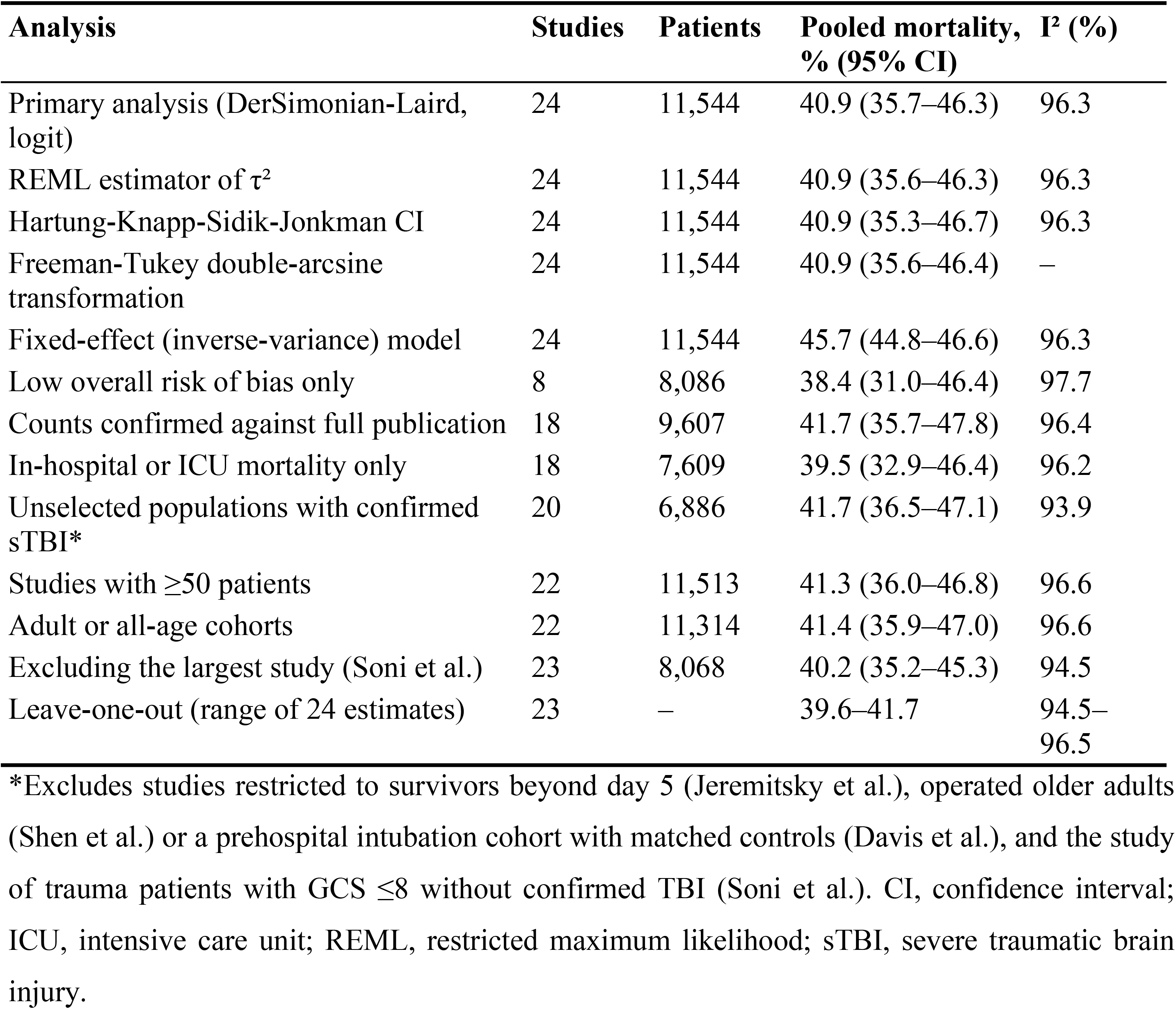
Sensitivity analyses.

#### Influence analyses

Omitting one study at a time gave estimates between 39.6% and 41.7% (Fig 6; S1 Table). The two studies with the highest mortality [47,49] had standardised residuals greater than 1.96; omitting either changed the pooled estimate by 1.3 percentage points.

**Fig 6.**
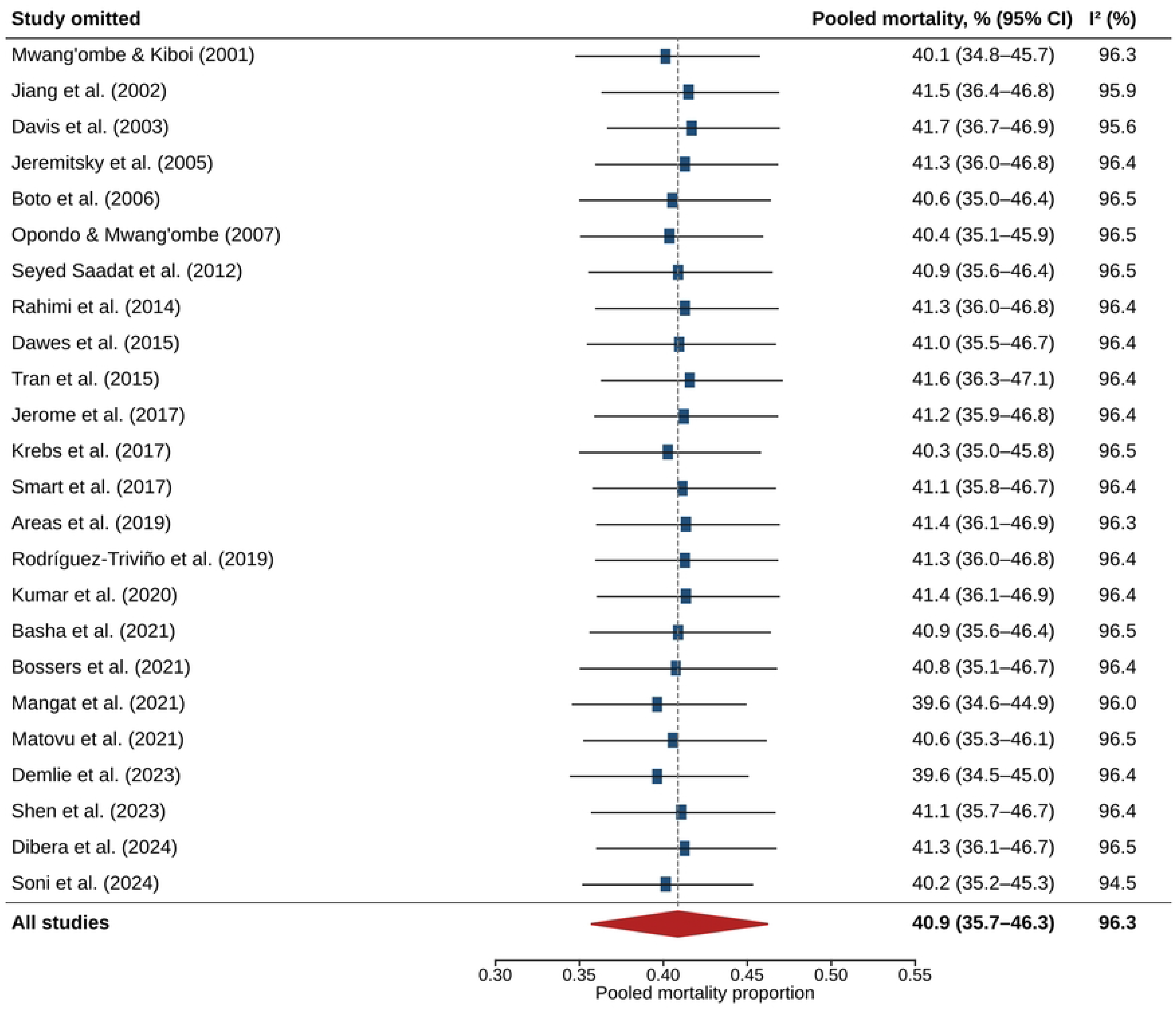
Leave-one-out sensitivity analysis. Each row shows the pooled random-effects mortality (95% CI) after omission of the named study.

#### Restricted analyses

Restricting the analysis to studies with low risk of bias (38.4%), studies with confirmed counts (41.7%), in-hospital or ICU mortality (39.5%), unselected populations with confirmed severe TBI (41.7%), studies with ≥50 patients (41.3%) or adult and all-age cohorts (41.4%) did not materially change the results. Heterogeneity remained considerable in all analyses (I² 93.9– 97.7%).

#### Small-study effects

The funnel plot showed no clear asymmetry, and Egger’s test did not indicate significant small-study effects (intercept −3.06, p = 0.09) (Fig 7). Given the high heterogeneity and the limitations of these methods for proportions, publication bias cannot be excluded.

**Fig 7.**
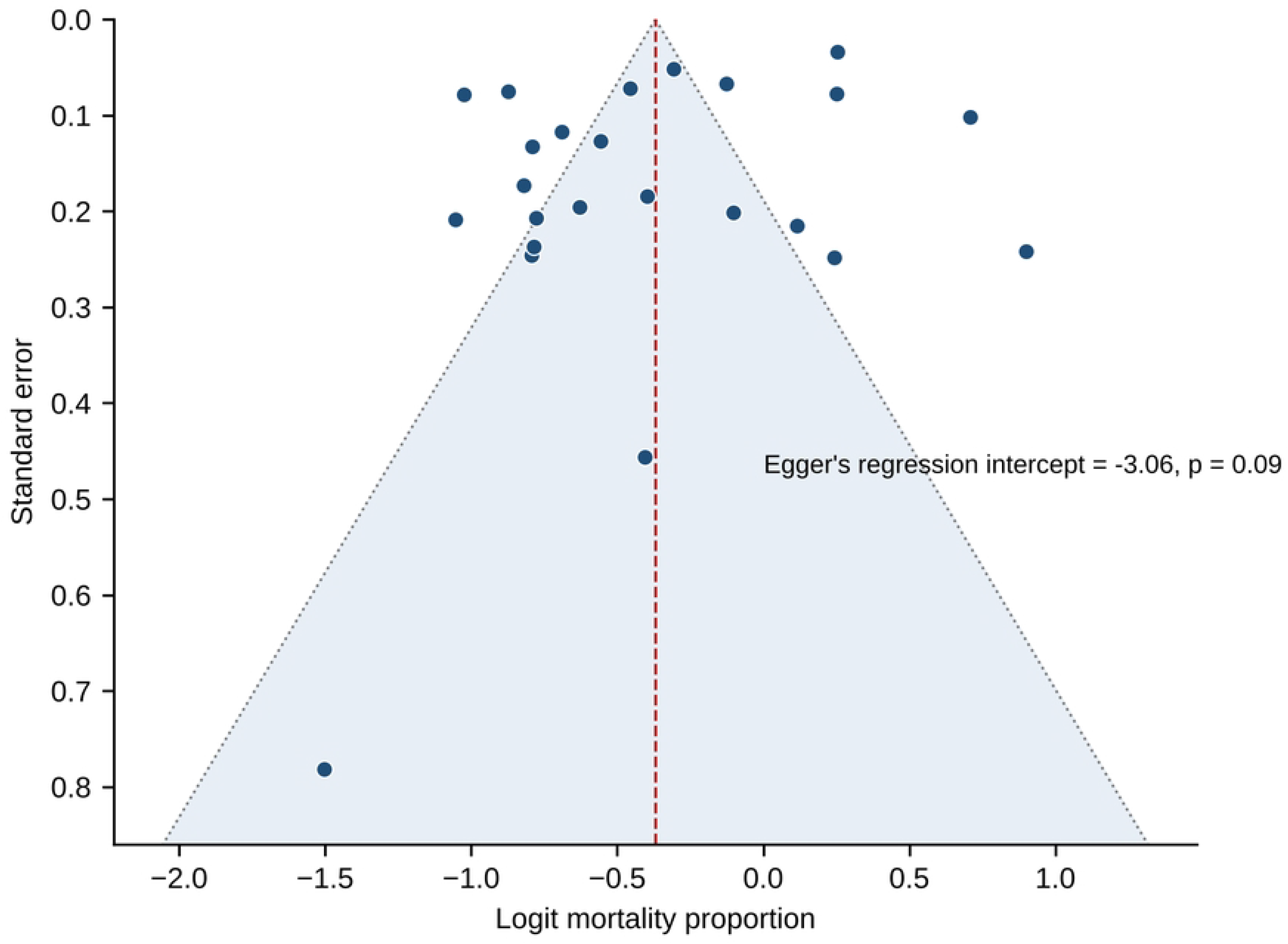
Funnel plot of logit mortality against its standard error. The dashed line shows the pooled logit estimate and the shaded region the pseudo-95% confidence limits.

#### Certainty of evidence

Certainty in the pooled mortality estimate was low (Table 5). Starting from high certainty for prognostic cohort evidence, it was downgraded one level each for:

- Risk of bias: two-thirds of studies had moderate risk, mostly from incomplete description of case mix.
- Inconsistency: I² above 90% and a prediction interval spanning roughly 19% to 67%, only partly explained by region.
- Indirectness: variation in GCS timing and definition, different outcome time points, and inclusion of selected populations.

The estimate was not downgraded for imprecision, because the 95% CI was narrow relative to the prediction interval, or for publication bias.

**Table 5.**
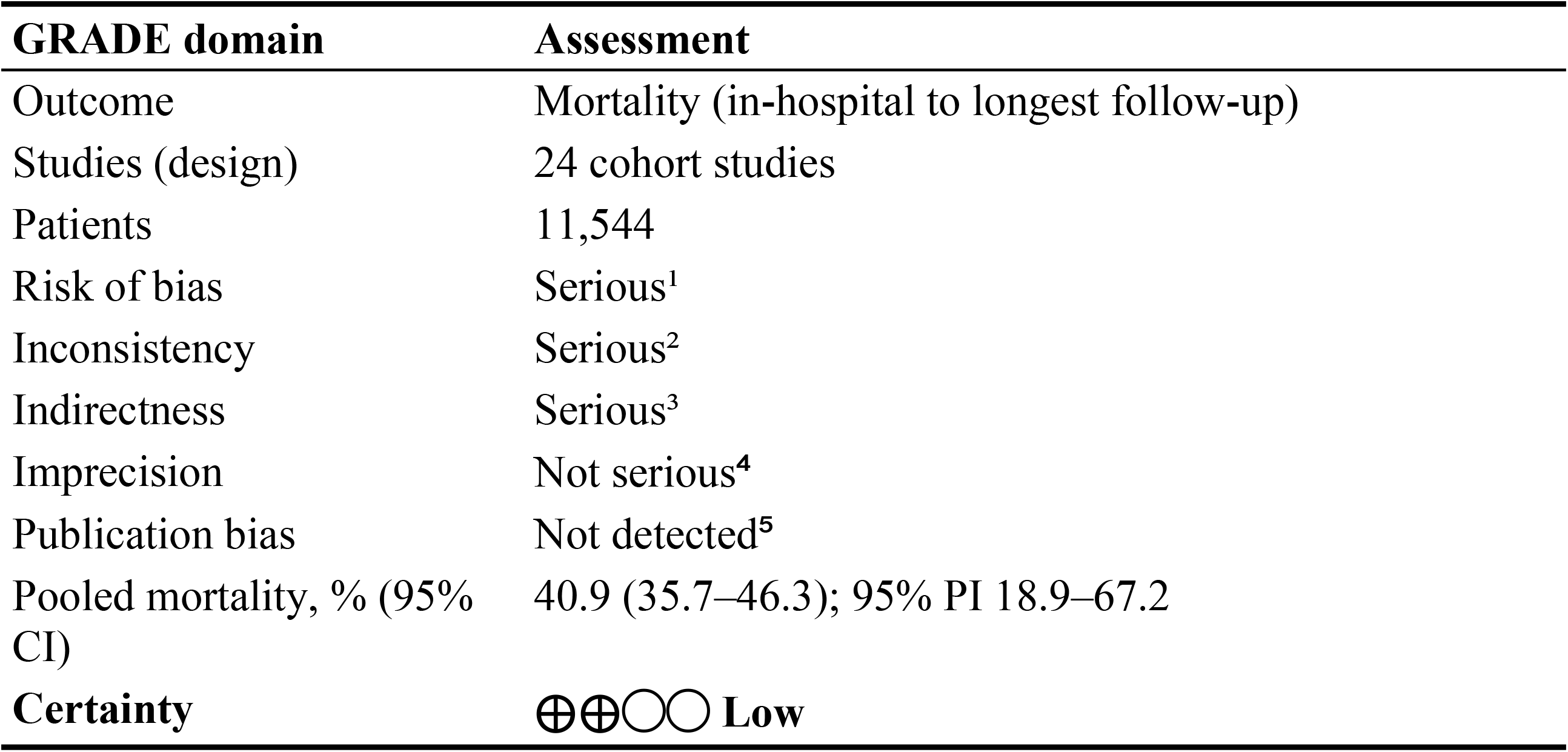
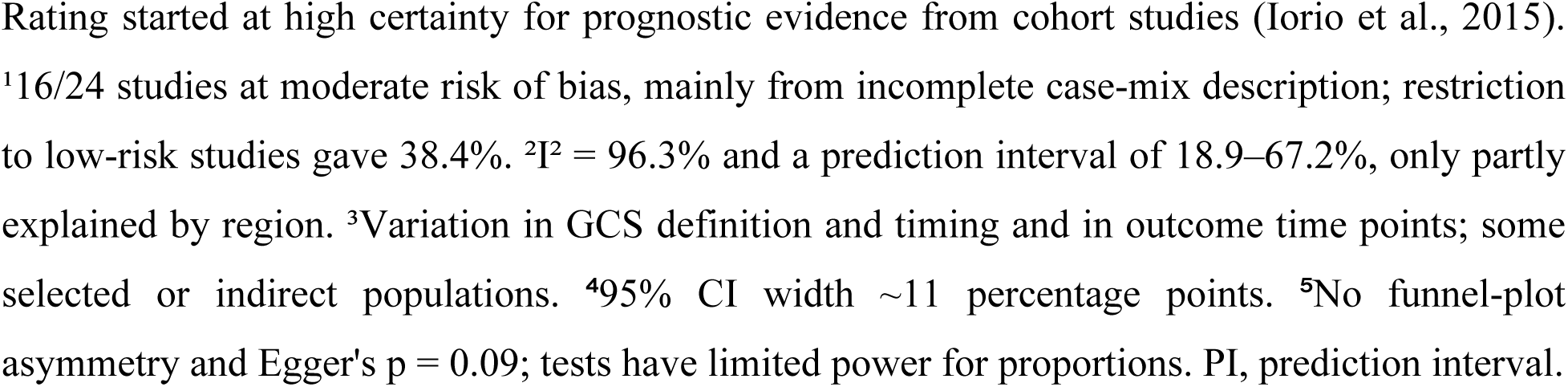
GRADE assessment of certainty for the pooled mortality estimate.

| GRADE domain | Assessment |
| --- | --- |
| Outcome | Mortality (in-hospital to longest follow-up) |
| Studies (design) | 24 cohort studies |
| Patients | 11,544 |
| Risk of bias | Serious<sup>1</sup> |
| Inconsistency | Serious<sup>2</sup> |
| Indirectness | Serious<sup>3</sup> |
| Imprecision | Not serious<sup>4</sup> |
| Publication bias | Not detected<sup>5</sup> |
| Pooled mortality, % (95% CI) | 40.9 (35.7–46.3); 95% PI 18.9–67.2 |
| Certainty | ⊕⊕○○ Low |

Rating started at high certainty for prognostic evidence from cohort studies (Iorio et al., 2015). <sup>1</sup>16/24 studies at moderate risk of bias, mainly from incomplete case-mix description; restriction to low-risk studies gave 38.4%. <sup>2</sup> $I^2 = 96.3\%$ and a prediction interval of 18.9–67.2%, only partly explained by region. <sup>3</sup>Variation in GCS definition and timing and in outcome time points; some selected or indirect populations. <sup>4</sup>95% CI width ~11 percentage points. <sup>5</sup>No funnel-plot asymmetry and Egger's $p = 0.09$ ; tests have limited power for proportions. PI, prediction interval.

## Discussion

### Principal findings

In this systematic review of 24 studies including 11,544 patients, approximately two in five patients with severe TBI died (pooled mortality 40.9%). Heterogeneity was considerable, and the prediction interval of roughly 19% to 67% is arguably the more clinically informative result: it indicates that the mortality expected in a given hospital or trauma system may be anywhere from one in five to two in three.

Mortality was highest in African cohorts and lowest in cohorts from the Americas, whereas differences by country income group, outcome time point, age group and publication year were not statistically significant. The pooled estimate was robust to the choice of estimator, to the omission of any single study and to restriction to studies at low risk of bias or with confirmed data.

### Comparison with previous evidence

Our pooled estimate is consistent with the long-term analysis by Stein and colleagues, who described a decline in severe-TBI mortality during the twentieth century followed by a plateau [5], and with the absence of a clear time trend among the studies included here (2001–2024).

The higher mortality in African cohorts is in keeping with multinational data showing worse outcomes in LMICs than in HICs after adjustment for prognostic factors [7,12], and with the Global Neurotrauma Outcomes Study, in which mortality after emergency neurosurgery was higher in countries with a lower human development index [8]. Contemporary HIC cohorts such as CENTER-TBI illustrate how case mix and care pathways vary even between well-resourced centres [53].

### Regional and income differences

The regional gradient did not translate into a significant difference by income group. Only six HIC studies were available, and they were heterogeneous: one was a prehospital cohort including patients with GCS ≤8 at the scene [46], and another was a prehospital intubation study with matched controls [31]. Conversely, some LMIC cohorts, such as patients admitted to a surgical ward [51] or registry-based audits [39], may have under-ascertained early deaths in the emergency department or before admission.

Region and income are therefore best regarded as markers of differences in trauma-system organisation, prehospital care, access to computed tomography and neurosurgery, ICU capacity and treatment-limitation practices, rather than as causes in themselves [54].

### Timing of mortality assessment

Mortality at different time points should not be treated as interchangeable. Deaths occurring before 48 hours mainly reflect the severity of the primary injury, whereas later deaths reflect secondary injury, complications and decisions on the continuation of care [10]. In the one included study reporting both, 19.8% of patients died within 48 hours and 46.8% died overall [33].

Because most studies reported only in-hospital mortality, and only two studies contributed to each other category, our data cannot establish whether mortality rises with longer follow-up. Future studies should report mortality at standardised time points, ideally 14 days (as in the CRASH models) and 6 months (as in IMPACT), in addition to in-hospital mortality [12,13].

### Implications for practice, policy and research

#### Clinical practice and trauma systems

A pooled mortality of about 40%, with a wide plausible range, provides a benchmark against which trauma systems can audit their outcomes, ideally after adjustment for case mix using validated prognostic models [12,13]. The high mortality observed in several African cohorts supports investment in prehospital care and timely transfer, access to computed tomography and neurosurgery, ICU capacity and adherence to evidence-based management guidelines [4,41,47].

### Future research

Research priorities include prospective multicentre registries in LMICs that use a uniform definition of severe TBI (GCS ≤8 after resuscitation), report deaths at fixed time points with complete follow-up, and record core case-mix variables. Such data would allow differences in outcome to be attributed to care rather than to case mix [54,55].

### Strengths and limitations

#### Strengths

Strengths of this review include a comprehensive search of six sources, duplicate study selection and data extraction, re-verification of extracted counts against the original publications, and a pre-specified hierarchy that avoided double counting of patients. The use of the logit transformation with prediction intervals and an extensive set of sensitivity analyses consistently supported the main estimate.

#### Limitations

Several limitations should be considered:

- Heterogeneity was considerable and largely unexplained, and subgroup analyses were observational comparisons between studies, susceptible to confounding.
- The definition and timing of the GCS assessment varied (for example, prehospital GCS, GCS after resuscitation or GCS <8), and outcome time points differed.
- Some populations were selected (survivors beyond day 5, operated older adults, prehospital intubation cohorts) or indirect (trauma patients with GCS ≤8 without confirmed TBI), although excluding them did not change the results.
- Evidence from Europe and from paediatric populations was limited to two studies each.
- The Newcastle-Ottawa Scale was adapted for single-group prognostic studies, and most studies did not describe case mix in enough detail to allow risk adjustment.
- Publication bias and selective reporting cannot be excluded, and individual-patient data were not available.

## Conclusions

Approximately two in five patients with severe TBI die, but expected mortality varies widely between settings, from about one in five to two in three, and is highest in African cohorts. Pooled estimates should therefore be used as a broad benchmark rather than as an individual risk.

Standardised definitions, fixed outcome time points and case-mix reporting are needed to allow valid comparisons between trauma systems and to identify modifiable causes of excess mortality.

## Acknowledgments

The authors thank their families for their support and encouragement throughout this work, and give thanks to God for strength and guidance.

## Author contributions

Conceptualization: FJR, BM, MM, TH, OE. Data curation: FJR, BM, OE. Formal analysis: FJR, MM. Investigation: FJR, BM, TH, OE. Methodology: FJR, BM, MM, TH. Project administration: FJR, BM. Resources: BM, MM, TH. Software: FJR. Supervision: BM, MM, TH. Validation: FJR, BM, MM. Visualization: FJR, OE. Writing – original draft: FJR. Writing – review & editing: all authors.

## Funding

The authors received no specific funding for this work.

## Competing interests

The authors have declared that no competing interests exist.

## Data availability

All data underlying the findings are within the manuscript and its Supporting Information files. The study-level dataset (S1 Dataset) and analysis code (S1 Code) are provided; the original data are available in the cited publications.

## Supporting information

**S1 Checklist.** PRISMA 2020 checklist.

**S1 File.** Full electronic search strategies for each database, with dates of the final searches.

**S1 Table.** Leave-one-out sensitivity analysis: pooled mortality, 95% CI and I² after omission of each study.

**S1 Dataset.** Study-level extracted data used in all analyses.

**S1 Code.** Python code used to perform the meta-analyses and produce the figures.

## Notes

### Competing Interest Statement

The authors have declared no competing interest.

### Clinical Protocols

https://www.crd.york.ac.uk/PROSPEROFILES/84de1c9c14d47a5f0dc515c79f0d1860.pdf.

### Author Declarations

Not applicable. This study is a systematic review and meta-analysis based exclusively on aggregate data from previously published studies. No individual participant-level data, human specimens, or tissue were collected or accessed by the authors therefore, institutional review board/ethics committee approval was not required.

## References

1. GBD 2016 Traumatic Brain Injury and Spinal Cord Injury Collaborators. Global, regional, and national burden of traumatic brain injury and spinal cord injury, 1990–2016: a systematic analysis for the Global Burden of Disease Study 2016. Lancet Neurol. 2019;18(1):56–87. doi: 10.1016/S1474-4422(18)30415-0

2. Dewan MC, Rattani A, Gupta S, Baticulon RE, Hung YC, Punchak M, et al. Estimating the global incidence of traumatic brain injury. J Neurosurg. 2019;130(4):1080–1097. doi: 10.3171/2017.10.JNS17352

3. Maas AIR, Menon DK, Manley GT, Abrams M, Åkerlund C, Andelic N, et al. Traumatic brain injury: progress and challenges in prevention, clinical care, and research. Lancet Neurol. 2022;21(11):1004–1060. doi: 10.1016/S1474-4422(22)00309-X

4. Carney N, Totten AM, O’Reilly C, Ullman JS, Hawryluk GWJ, Bell MJ, et al. Guidelines for the management of severe traumatic brain injury, fourth edition. Neurosurgery. 2017;80(1):6–15. doi: 10.1227/NEU.0000000000001432

5. Stein SC, Georgoff P, Meghan S, Mizra K, Sonnad SS. 150 years of treating severe traumatic brain injury: a systematic review of progress in mortality. J Neurotrauma. 2010;27(7):1343–1353. doi: 10.1089/neu.2009.1206

6. Georgoff P, Meghan S, Mirza K, Stein SC. Geographic variation in outcomes from severe traumatic brain injury. World Neurosurg. 2010;74(2–3):331–345. doi: 10.1016/j.wneu.2010.03.025

7. De Silva MJ, Roberts I, Perel P, Edwards P, Kenward MG, Fernandes J, et al. Patient outcome after traumatic brain injury in high-, middle- and low-income countries: analysis of data on 8927 patients in 46 countries. Int J Epidemiol. 2009;38(2):452–458. doi: 10.1093/ije/dyn189

8. Clark D, Joannides A, Adeleye AO, Bajamal AH, Bashford T, Biluts H, et al. Casemix, management, and mortality of patients receiving emergency neurosurgery for traumatic brain injury in the Global Neurotrauma Outcomes Study: a prospective observational cohort study. Lancet Neurol. 2022;21(5):438–449. doi: 10.1016/S1474-4422(22)00037-0

9. Mauritz W, Wilbacher I, Majdan M, Leitgeb J, Janciak I, Brazinova A, et al. Epidemiology, treatment and outcome of patients after severe traumatic brain injury in European regions with different economic status. Eur J Public Health. 2008;18(6):575–580. doi: 10.1093/eurpub/ckn079

10. Tisherman SA, Schmicker RH, Brasel KJ, Bulger EM, Kerby JD, Minei JP, et al. Detailed description of all deaths in both the shock and traumatic brain injury hypertonic saline trials of the Resuscitation Outcomes Consortium. Ann Surg. 2015;261(3):586–590. doi: 10.1097/SLA.0000000000000837

11. Roozenbeek B, Chiu YL, Lingsma HF, Gerber LM, Steyerberg EW, Ghajar J, et al. Predicting 14-day mortality after severe traumatic brain injury: application of the IMPACT models in the Brain Trauma Foundation TBI-trac New York State database. J Neurotrauma. 2012;29(7):1306–1312. doi: 10.1089/neu.2011.1988

12. MRC CRASH Trial Collaborators. Predicting outcome after traumatic brain injury: practical prognostic models based on large cohort of international patients. BMJ. 2008;336(7641):425–429. doi: 10.1136/bmj.39461.643438.25

13. Steyerberg EW, Mushkudiani N, Perel P, Butcher I, Lu J, McHugh GS, et al. Predicting outcome after traumatic brain injury: development and international validation of prognostic scores based on admission characteristics. PLoS Med. 2008;5(8):e165. doi: 10.1371/journal.pmed.0050165

14. Page MJ, McKenzie JE, Bossuyt PM, Boutron I, Hoffmann TC, Mulrow CD, et al. The PRISMA 2020 statement: an updated guideline for reporting systematic reviews. BMJ. 2021;372:n71. doi: 10.1136/bmj.n71

15. Munn Z, Moola S, Lisy K, Riitano D, Tufanaru C. Methodological guidance for systematic reviews of observational epidemiological studies reporting prevalence and cumulative incidence data. Int J Evid Based Healthc. 2015;13(3):147–153. doi: 10.1097/XEB.0000000000000054

16. Wells GA, Shea B, O’Connell D, Peterson J, Welch V, Losos M, et al. The Newcastle-Ottawa Scale (NOS) for assessing the quality of nonrandomised studies in meta-analyses. Ottawa: Ottawa Hospital Research Institute; 2000. Available from: https://www.ohri.ca/programs/clinical_epidemiology/oxford.asp

17. DerSimonian R, Laird N. Meta-analysis in clinical trials. Control Clin Trials. 1986;7(3):177–188. doi: 10.1016/0197-2456(86)90046-2

18. Barendregt JJ, Doi SA, Lee YY, Norman RE, Vos T. Meta-analysis of prevalence. J Epidemiol Community Health. 2013;67(11):974–978. doi: 10.1136/jech-2013-203104

19. Schwarzer G, Chemaitelly H, Abu-Raddad LJ, Rücker G. Seriously misleading results using inverse of Freeman-Tukey double arcsine transformation in meta-analysis of single proportions. Res Synth Methods. 2019;10(3):476–483. doi: 10.1002/jrsm.1348

20. Higgins JPT, Thompson SG, Deeks JJ, Altman DG. Measuring inconsistency in meta-analyses. BMJ. 2003;327(7414):557–560. doi: 10.1136/bmj.327.7414.557

21. Riley RD, Higgins JPT, Deeks JJ. Interpretation of random effects meta-analyses. BMJ. 2011;342:d549. doi: 10.1136/bmj.d549

22. Higgins JPT, Thomas J, Chandler J, Cumpston M, Li T, Page MJ, et al., editors. Cochrane Handbook for Systematic Reviews of Interventions. Version 6.5. Cochrane; 2024. Available from: https://www.training.cochrane.org/handbook

23. IntHout J, Ioannidis JPA, Borm GF. The Hartung-Knapp-Sidik-Jonkman method for random effects meta-analysis is straightforward and considerably outperforms the standard DerSimonian-Laird method. BMC Med Res Methodol. 2014;14:25. doi: 10.1186/1471-2288-14-25

24. Viechtbauer W, Cheung MWL. Outlier and influence diagnostics for meta-analysis. Res Synth Methods. 2010;1(2):112–125. doi: 10.1002/jrsm.11

25. Egger M, Davey Smith G, Schneider M, Minder C. Bias in meta-analysis detected by a simple, graphical test. BMJ. 1997;315(7109):629–634. doi: 10.1136/bmj.315.7109.629

26. Hunter JP, Saratzis A, Sutton AJ, Boucher RH, Sayers RD, Bown MJ. In meta-analyses of proportion studies, funnel plots were found to be an inaccurate method of assessing publication bias. J Clin Epidemiol. 2014;67(8):897–903. doi: 10.1016/j.jclinepi.2014.03.003

27. Iorio A, Spencer FA, Falavigna M, Alba C, Lang E, Burnand B, et al. Use of GRADE for assessment of evidence about prognosis: rating confidence in estimates of event rates in broad categories of patients. BMJ. 2015;350:h870. doi: 10.1136/bmj.h870

28. Lanzillo B, Piscosquito G, Marcuccio L, Lanzillo A, Vitale DF. Prognosis of severe acquired brain injury: short and long-term outcome determinants and their potential clinical relevance after rehabilitation. A comprehensive approach to analyze cohort studies. PLoS One. 2019;14(9):e0216507. doi: 10.1371/journal.pone.0216507

29. Mwang’ombe NJM, Kiboi J. Factors influencing the outcome of severe head injury at Kenyatta National Hospital. East Afr Med J. 2001;78(5):238–241. doi: 10.4314/eamj.v78i5.9045

30. Jiang JY, Gao GY, Li WP, Yu MK, Zhu C. Early indicators of prognosis in 846 cases of severe traumatic brain injury. J Neurotrauma. 2002;19(7):869–874. doi: 10.1089/08977150260190456

31. Davis DP, Hoyt DB, Ochs M, Fortlage D, Holbrook T, Marshall LK, et al. The effect of paramedic rapid sequence intubation on outcome in patients with severe traumatic brain injury. J Trauma. 2003;54(3):444–453. doi: 10.1097/01.TA.0000053396.02126.CD

32. Jeremitsky E, Omert LA, Dunham CM, Wilberger J, Rodriguez A. The impact of hyperglycemia on patients with severe brain injury. J Trauma. 2005;58(1):47–50. doi: 10.1097/01.TA.0000135158.42242.B1

33. Boto GR, Gómez PA, De La Cruz J, Lobato RD. Severe head injury and the risk of early death. J Neurol Neurosurg Psychiatry. 2006;77(9):1054–1059. doi: 10.1136/jnnp.2005.087056

34. Opondo EA, Mwang’ombe NJM. Outcome of severe traumatic brain injury at a critical care unit: a review of 87 patients. Ann Afr Surg. 2007;1:3–9. doi: 10.4314/aas.v1i1.45788

35. Seyed Saadat SM, Bidabadi E, Seyed Saadat SN, Mashouf M, Salamat F, Yousefzadeh S. Association of persistent hyperglycemia with outcome of severe traumatic brain injury in pediatric population. Childs Nerv Syst. 2012;28(10):1773–1777. doi: 10.1007/s00381-012-1753-5

36. Rahimi S, Bidabadi E, Mashouf M, Seyed Saadat SM, Rahimi S. Prognostic value of arterial blood gas disturbances for in-hospital mortality in pediatric patients with severe traumatic brain injury. Acta Neurochir (Wien). 2014;156(1):187–192. doi: 10.1007/s00701-013-1929-0

37. Dawes AJ, Sacks GD, Cryer HG, Gruen JP, Preston C, Gorospe D, et al. Intracranial pressure monitoring and inpatient mortality in severe traumatic brain injury: a propensity score-matched analysis. J Trauma Acute Care Surg. 2015;78(3):492–501. doi: 10.1097/TA.0000000000000559

38. Tran TM, Fuller AT, Kiryabwire J, Mukasa J, Muhumuza M, Ssenyojo H, et al. Distribution and characteristics of severe traumatic brain injury at Mulago National Referral Hospital in Uganda. World Neurosurg. 2015;83(3):269–277. doi: 10.1016/j.wneu.2014.12.028

39. Jerome E, Laing GL, Bruce JL, Sartorius B, Brysiewicz P, Clarke DL. An audit of traumatic brain injury (TBI) in a busy developing-world trauma service exposes a significant deficit in resources available to manage severe TBI. S Afr Med J. 2017;107(7):621–625. doi: 10.7196/SAMJ.2017.v107i7.10562

40. Krebs E, Gerardo CJ, Park LP, Vissoci JRN, Byiringiro JC, Byiringiro F, et al. Mortality-associated characteristics of patients with traumatic brain injury at the University Teaching Hospital of Kigali, Rwanda. World Neurosurg. 2017;102:571–582. doi: 10.1016/j.wneu.2017.03.001

41. Smart LR, Mangat HS, Issarow B, McClelland P, Mayaya G, Kanumba E, et al. Severe traumatic brain injury at a tertiary referral center in Tanzania: epidemiology and adherence to Brain Trauma Foundation guidelines. World Neurosurg. 2017;105:238–248. doi: 10.1016/j.wneu.2017.05.101

42. Areas FZ, Schwarzbold ML, Diaz AP, Rodrigues IK, Sousa DS, Ferreira CL, et al. Predictors of hospital mortality and the related burden of disease in severe traumatic brain injury: a prospective multicentric study in Brazil. Front Neurol. 2019;10:432. doi: 10.3389/fneur.2019.00432

43. Rodríguez-Triviño CY, Torres Castro I, Dueñas Z. Hypochloremia in patients with severe traumatic brain injury: a possible risk factor for increased mortality. World Neurosurg. 2019;124:e783–e788. doi: 10.1016/j.wneu.2019.01.025

44. Kumar RG, DiSanto D, Awan N, Vaughan LE, Levochkina MS, Weppner JL, et al. Temporal acute serum estradiol and tumor necrosis factor-α associations and risk of death after severe traumatic brain injury. J Neurotrauma. 2020;37(20):2198–2210. doi: 10.1089/neu.2019.6577

45. Basha AK, Mahmoud MA, Al Ashwal MI, Aglan O, ElShawady SB, Abdel-Latif AM, et al. Management of severe traumatic brain injury: a single institution experience in a middle-income country. Front Surg. 2021;8:690723. doi: 10.3389/fsurg.2021.690723

46. Bossers SM, Boer C, Bloemers FW, Van Lieshout EMM, Den Hartog D, Hoogerwerf N, et al. Epidemiology, prehospital characteristics and outcomes of severe traumatic brain injury in the Netherlands: the BRAIN-PROTECT study. Prehosp Emerg Care. 2021;25(5):644–655. doi: 10.1080/10903127.2020.1824049

47. Mangat HS, Wu X, Gerber LM, Shabani HK, Lazaro A, Leidinger A, et al. Severe traumatic brain injury management in Tanzania: analysis of a prospective cohort. J Neurosurg. 2021;135(4):1190–1202. doi: 10.3171/2020.8.JNS201243

48. Matovu P, Kirya M, Galukande M, Kiryabwire J, Mukisa J, Ocen W, et al. Hyperglycemia in severe traumatic brain injury patients and its association with thirty-day mortality: a prospective observational cohort study in Uganda. PeerJ. 2021;9:e10589. doi: 10.7717/peerj.10589

49. Demlie TA, Alemu MT, Messelu MA, Wagnew F, Mekonen EG. Incidence and predictors of mortality among traumatic brain injury patients admitted to Amhara region Comprehensive Specialized Hospitals, northwest Ethiopia, 2022. BMC Emerg Med. 2023;23:55. doi: 10.1186/s12873-023-00823-9

50. Shen H, Liu H, He J, Wei L, Wang S. Risk factors of prognosis in older patients with severe brain injury after surgical intervention. Eur J Med Res. 2023;28:479. doi: 10.1186/s40001-023-01473-0

51. Dibera GB, Yizengaw MA, Yadeta GL, Iticha DD, Gamachu B, Legesse BT. Clinical characteristics and outcomes of traumatic brain injury in patients admitted to surgical ward of Jimma Medical Center, Southwest Ethiopia: a prospective observational follow-up study. BMJ Open. 2024;14(2):e080598. doi: 10.1136/bmjopen-2023-080598

52. Soni KD, Bansal V, Khajanchi M, Kizhakke Veetil D, Anderson G, Rayker N, et al. Intubation and in-hospital mortality after trauma with Glasgow Coma Scale score eight or less: a cohort study. J Surg Res. 2024;299:188–194. doi: 10.1016/j.jss.2024.04.014

53. Steyerberg EW, Wiegers E, Sewalt C, Buki A, Citerio G, De Keyser V, et al. Case-mix, care pathways, and outcomes in patients with traumatic brain injury in CENTER-TBI: a European prospective, multicentre, longitudinal, cohort study. Lancet Neurol. 2019;18(10):923–934. doi: 10.1016/S1474-4422(19)30232-7

54. Rubiano AM, Carney N, Chesnut R, Puyana JC. Global neurotrauma research challenges and opportunities. Nature. 2015;527(7578):S193–S197. doi: 10.1038/nature16035

55. Chesnut RM, Temkin N, Carney N, Dikmen S, Rondina C, Videtta W, et al. A trial of intracranial-pressure monitoring in traumatic brain injury. N Engl J Med. 2012;367(26):2471–2481. doi: 10.1056/NEJMoa1207363

